# Trends and Outcomes of Carotid Revascularization for Asymptomatic Carotid Stenosis Among End-Stage-Kidney-Disease Patients on Dialysis from 2010 to 2019

**DOI:** 10.1101/2025.09.28.25336858

**Authors:** Gokul Ramani, Wan-Chi Chan, Zafar Ali, Kunal Patel, Monil Majmundar, Rhythm Vasudeva, Akshaya Gadre, Cyrus Mugunti, Kartik Munshi, Axel Thors, Seth DeCamp, Kamal Gupta, Gaurav Parmar

**Author notes:** CORRESPONDING AUTHOR: Gokul Ramani, MD, Address: 4000 Cambridge Blvd, Kansas City, KS 66160. Contributed equally (Gokul Ramani, Wan-Chi Chan).

## Abstract

**INTRODUCTION:** The role of carotid revascularization for asymptomatic carotid artery stenosis (ACAS) in dialysis-dependent end-stage kidney disease (ESKD) patients remains poorly defined, as these patients have high periprocedural risks and limited long-term survival. This study evaluated the trends in carotid revascularization in this group and studied associated short- and long-term outcomes.

**METHODS:** We analyzed the United States Renal Data System (USRDS) to study dialysis-dependent ESKD patients with ACAS who underwent carotid endarterectomy (CEA) or carotid artery stenosis (CAS) between 2010 and 2019. Primary outcomes included trends in CEA and CAS utilization, 30-day stroke or death rates. Secondary outcomes were in-hospital and one-year stroke or death rates.

**RESULTS:** Among 11,405 ESKD patients on dialysis with ACAS, 4954 underwent carotid revascularization (4098 CEA; 856 CAS). CEA rates reduced by 55% and CAS by 50% between 2010 and 2019.

CAS was associated with a higher 30-day composite stroke or death rate (7.13% vs 4.5%; P=0.0014), in-hospital stroke or death rate (3.39% vs 2.22%, P=0.0433), and 1-year stroke or death rate (33.1% vs 25.4%, P<0.001) compared to CEA. No significant improvements in the outcomes over time were observed.

**CONCLUSION:** Carotid revascularization rates for ACAS have declined among dialysis-dependent ESKD patients, yet both CEA and CAS are associated with significant procedure-related stroke and death risk. These support a cautious approach and underscore the need for a more selective and individualized shared decision-making approach in this high-risk population.

## INTRODUCTION

Stroke remains a leading cause of mortality and disability in the United States (1), with patients who have end-stage kidney disease (ESKD) experiencing disproportionately worse outcomes and higher post-stroke mortality than the general population (2). In patients with asymptomatic carotid artery stenosis (ACAS), the clinical benefit of carotid revascularization has been increasingly questioned. Although randomized trials have demonstrated modest stroke risk reduction with revascularization in appropriately selected patients (3-5), these benefits are counterbalanced by procedural risks, including periprocedural stroke and death, especially in patients who have reduced life expectancy (6-8). Hence, according to most professional society guidelines, revascularization for ACAS is recommended only in patients with low procedural risk and an anticipated all-cause survival of at least 3-5 years (Class IIa, Level B) (6,7). However, these recommendations are based on data from clinical trials that largely excluded patients with ESKD on dialysis – a population known to carry markedly higher cardiovascular morbidity, procedural risk, and limited survival (9-16).

Despite these concerns, a subset of ESKD patients with ACAS continue to undergo carotid revascularization without information on whether it meaningfully improves outcomes. Prior observational studies have suggested poor periprocedural and long-term outcomes following carotid revascularization in dialysis-dependent patients (9, 10). Yet, trends and comparative outcomes have not been fully characterized over time.

To address this knowledge gap, we used a national database to evaluate trends in the use of carotid endarterectomy (CEA) and carotid artery stenting (CAS) in ESKD patients on dialysis with ACAS from 1999 to 2019 and to assess short—and long-term outcomes, including stroke and mortality.

## METHODS

### Data Source

We utilized data from the United States Renal Data System (USRDS), a national registry of all patients with end-stage kidney disease (ESKD) in the United States (17, 18). The database includes detailed information on demographics, dialysis modality, comorbidities, procedures, and outcomes. Our study is exempt from the institutional review board approval since it used de-identified data in adherence with the Health Insurance Portability and Accountability Act (19).

### Study Population

We identified adults (≥18 years) on maintenance dialysis for at least ≥90 days who underwent CEA or CAS for ACAS between January 1, 2010, and December 31, 2019. CEA and CAS were identified using validated ICD-9 and ICD-10 procedure codes (Supplement Tables 1-2). Patients were excluded if they had evidence of symptomatic carotid disease within 6 months before revascularization (e.g., stroke, transient ischemic attack (TIA), or amaurosis fugax) (Supplement Table 3), lacked Medicare coverage at the time of the index procedure, had undergone both the procedures-CEA and CAS during the index hospitalization, had incomplete demographic or clinical data, had received a kidney transplant before or during the index hospitalization, and if the patients resided outside the United States (Figure 1). Only the first qualifying revascularization was included per patient. Patients were followed until death, kidney transplantation, loss of Medicare coverage, or December 31, 2020.

**Table 1:** Demographics and patient characteristics of CEA and CAS in the United States between 2010 and 2019.

| Characteristic | CEA |  | CAS |  |
| --- | --- | --- | --- | --- |
|  | 4098 (n) |  | 856 (n) |  |
|  | (n) | (%) | (n) | (%) |
| Male gender | 2403 | 58.6% | 512 | 59.8% |
| Race |  |  |  |  |
| White | 2817 | 68.7% | 519 | 60.6% |
| Black | 648 | 15.8% | 180 | 21.0% |
| Asian/Other | 633 | 15.5% | 157 | 18.3% |
| Mean Age (years) | 69.7 | 9.5% | 70.2 | 9.1% |
| Age groups (years) |  |  |  |  |
| <50 | 104 | 2.5% | 16 | 1.9% |
| 50-64 | 1094 | 26.7% | 220 | 25.7% |
| 65-79 | 2312 | 56.4% | 495 | 57.8% |
| ≥80 | 588 | 14.4% | 125 | 14.6% |
| Dialysis Modality |  |  |  |  |
| Hemodialysis | 3625 | 88.5% | 774 | 90.4% |
| Peritoneal dialysis | 473 | 11.5% | 82 | 9.6% |
| Dialysis duration (years) |  |  |  |  |
| <2 | 1917 | 46.8% | 388 | 45.3% |
| 2-5 | 1691 | 41.3% | 354 | 41.4% |
| 6-10 | 425 | 10.4% | 95 | 11.1% |
| ≥11 | 65 | 1.6% | 19 | 2.2% |
| Comorbid conditions |  |  |  |  |
| CAD | 2785 | 68.0% | 628 | 73.4% |
| CHF | 1930 | 47.1% | 477 | 55.7% |
| COPD | 753 | 18.4% | 177 | 20.7% |
| CVA / TIA | 747 | 18.2% | 237 | 27.7% |
| Cancer | 430 | 10.5% | 81 | 9.5% |
| Diabetes Mellitus | 2887 | 70.5% | 619 | 72.3% |
| Dysrhythmia | 881 | 21.5% | 210 | 24.5% |
| Hypertension | 3938 | 96.1% | 828 | 96.7% |
| Liver disease | 54 | 1.3% | <11 | - |
| PAD | 1916 | 46.8% | 412 | 48.1% |
| Atrial fibrillation/flutter | 722 | 17.6% | 158 | 18.5% |
| Obesity | 340 | 8.3% | 75 | 8.8% |
| Valvular Disease | 381 | 9.3% | 95 | 11.1% |
| Tobacco use/Smoking | 1597 | 39.0% | 304 | 35.5% |
| Long-term use of anti-platelet/thrombotic | 515 | 12.6% | 160 | 18.7% |
| Long-term (current) use of aspirin | 892 | 21.8% | 198 | 23.1% |

USRDS: United States Renal Data System, ESKD: End Stage Kidney-Disease, CEA: Carotid Endarterectomy, CAS: Carotid Artery Stenting, CAD: Coronary Artery Disease, CHF: Congestive Heart Failure, COPD: Chronic Obstructive Pulmonary Disease, CVA: Cerebrovascular Disease, TIA: Transient Ischemic Attack, PAD: Peripheral Arterial Disease

**Figure 1:**
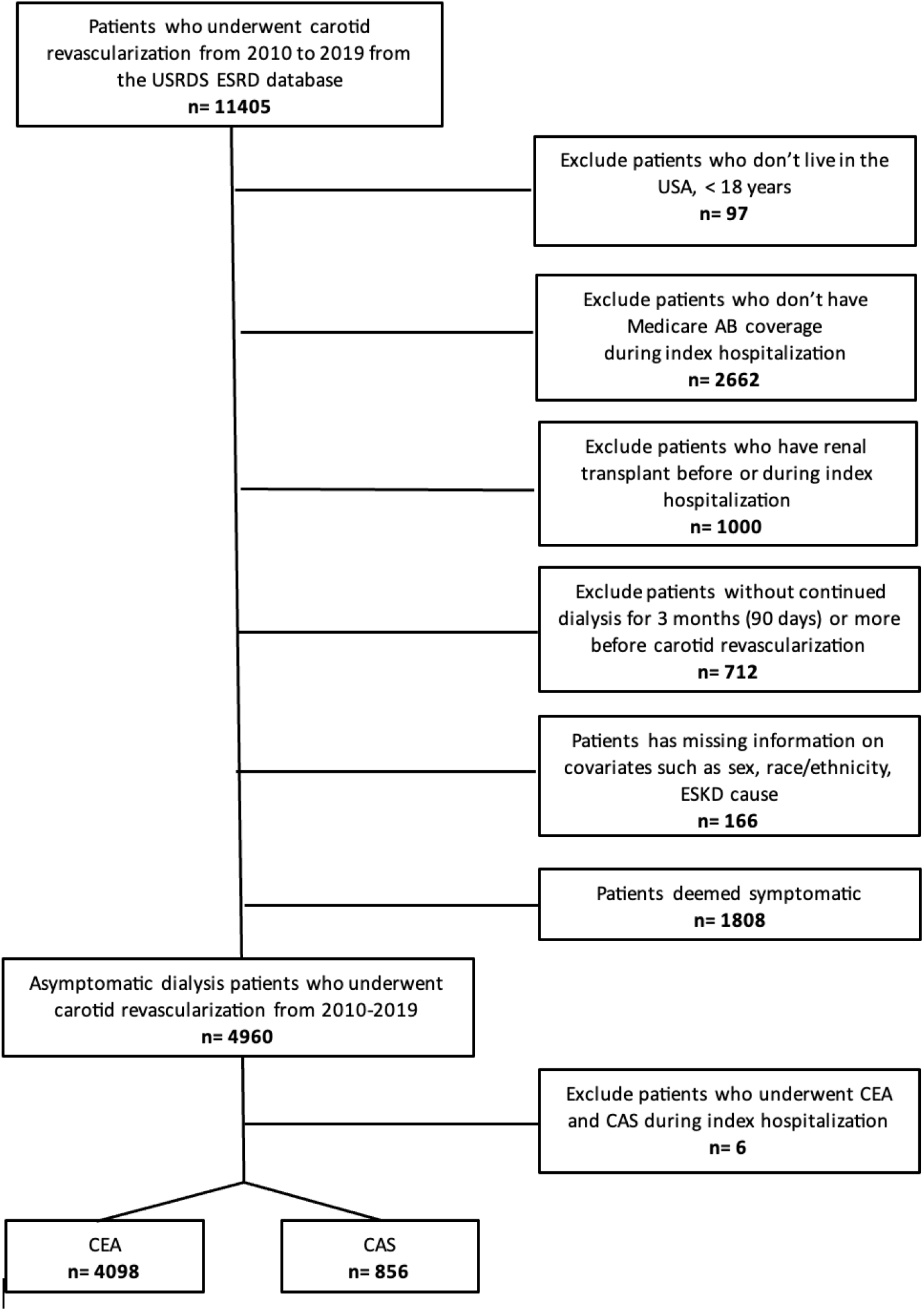
Study Population. USRDS: United States Renal Data System, ESKD: End Stage Kidney-Disease, CEA: Carotid Endarterectomy, CAS: Carotid Artery Stenting

### Covariates and Comorbidities

Baseline characteristics, including age, sex, race, ethnicity, dialysis modality and duration, and primary cause of ESKD, were obtained from CMS-2728 forms. Comorbid conditions (e.g., coronary artery disease (CAD), congestive heart failure (CHF), chronic obstructive pulmonary disease, atrial fibrillation, diabetes, and peripheral artery disease (PAD)) were ascertained from diagnosis codes in Medicare claims during the 12 months before the procedure.

### Outcomes

The primary outcomes were annual trends in CEA and CAS rates as well as the 30-day post-discharge composite of stroke or death. Secondary outcomes included in-hospital stroke or death and one-year stroke or death. Stroke was defined using validated ICD-9 and ICD-10 codes for ischemic stroke, hemiplegia, or cerebral infarction.

### Statistical Analysis

Categorical variables were summarized as frequencies (percentages) and compared using the Chi-squared test. Continuous variables were presented as means and standard deviations and compared using two-sample t-tests. Procedure rates were calculated per 100,000 patients with ESKD. Trends over time were visualized with line plots. Kaplan-Meier methods were used to estimate the 30-day and 1-year cumulative incidence of stroke or death, with comparisons by the long-rank test. All analyses were performed using SAS version 9.4 (SAS Institute Inc.). A 2-sided P value of <0.05 was considered statistically significant.

## RESULTS

### Study Population

A total of 11,405 ESKD patients on dialysis were identified as having ACAS in the USRDS from 2010 to 2019. Of these, 4954 patients underwent carotid revascularization and met the inclusion criteria: 4098 (83%) underwent CEA, and 856 (17%) underwent CAS (Figure 1). Baseline characteristics are depicted in Table 1. Patients undergoing CAS were slightly older (mean age, 70.2 ± 9.1 vs 69.7 ± 9.5 years for CEA) and more likely to be Black (21.0% vs 15.8%) or of Asian or other races (18.3% vs 15.5%). A higher proportion of CAS patients had prior stroke or TIA (27.7% vs 18.2%), although these events were experienced more than six months before the index procedure. The majority in both groups were on hemodialysis (CAS: 90.4%, CEA: 88.5%). The comorbidity burden was relatively high in both groups. CAS patients had a higher prevalence of CAD (73.4% vs 68.0%), CHF (55.7% vs 47.1%), and PAD (48.1% vs 46.8%).

### Trends in Procedure Utilization

Between 2010 and 2019, the overall use of carotid revascularization for ACAS among ESKD patients declined significantly. The rate of CEA decreased from 87 to 39 per 100,000 patients with ESKD (55% overall decline), while the CAS rate decreased from 22 to 11 per 100,000 (50% overall decline). On average, CEA procedures declined by 4.85% annually, and CAS by 3.82% annually (P for trend < 0.001 for both) (Figure 2).

**Figure 2:**
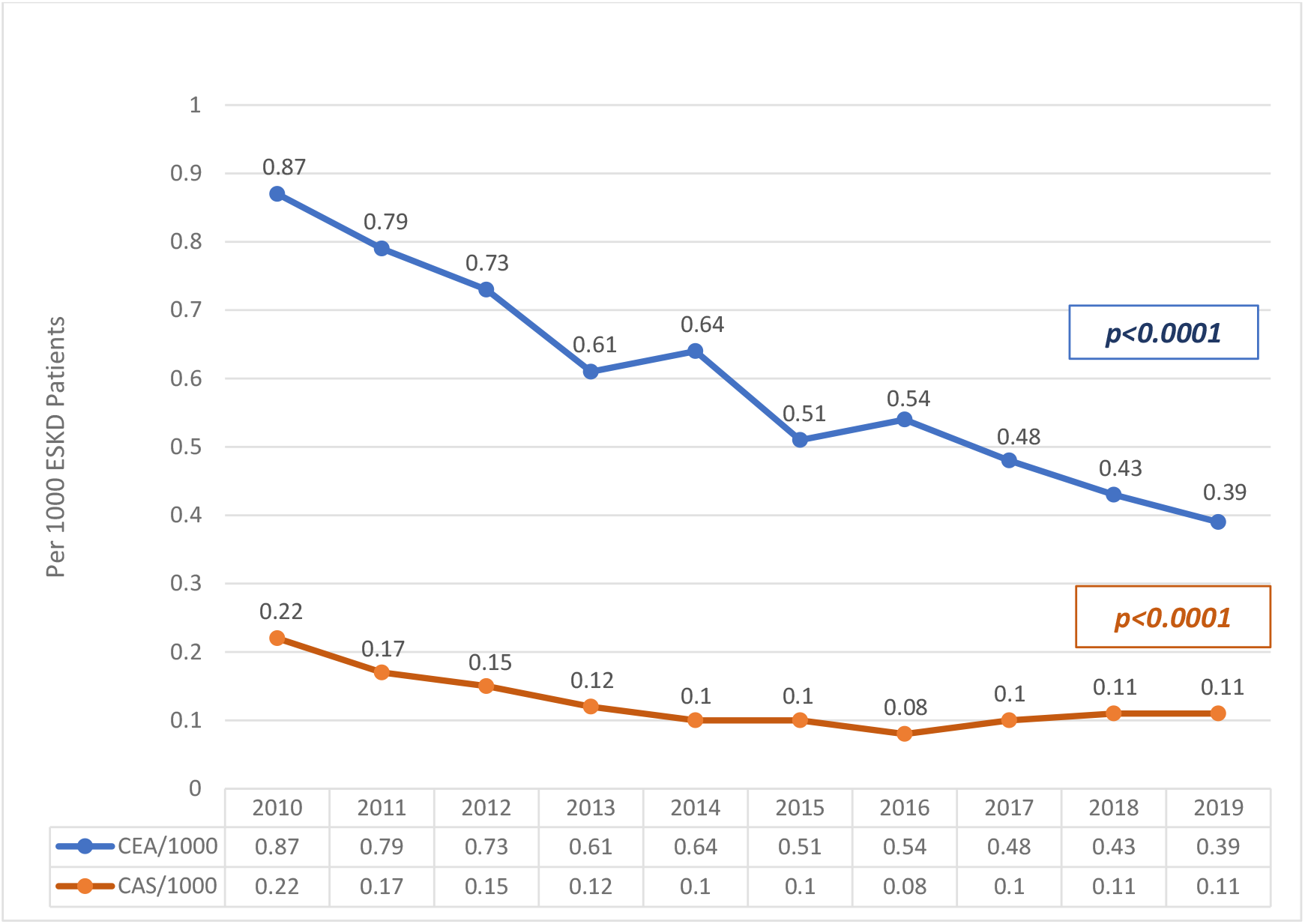
Annual trends in carotid revascularizations Performed for asymptomatic carotid stenosis per 1000 ESKD Patients from 2010-2019. CEA: Carotid Endarterectomy, CAS: Carotid Artery Stenting

### Thirty-Day Stroke or Death

The thirty-day post-discharge composite stroke or death did not significantly change over time for either procedure (P for trend: CEA=0.15; CAS=0.65) (Figure 3). However, comparing pooled data across the decade, the 30-day event rate was significantly higher for CAS than for CEA (7.13% vs 4.5%, P=0.0014) (Figure 5).

**Figure 3:**
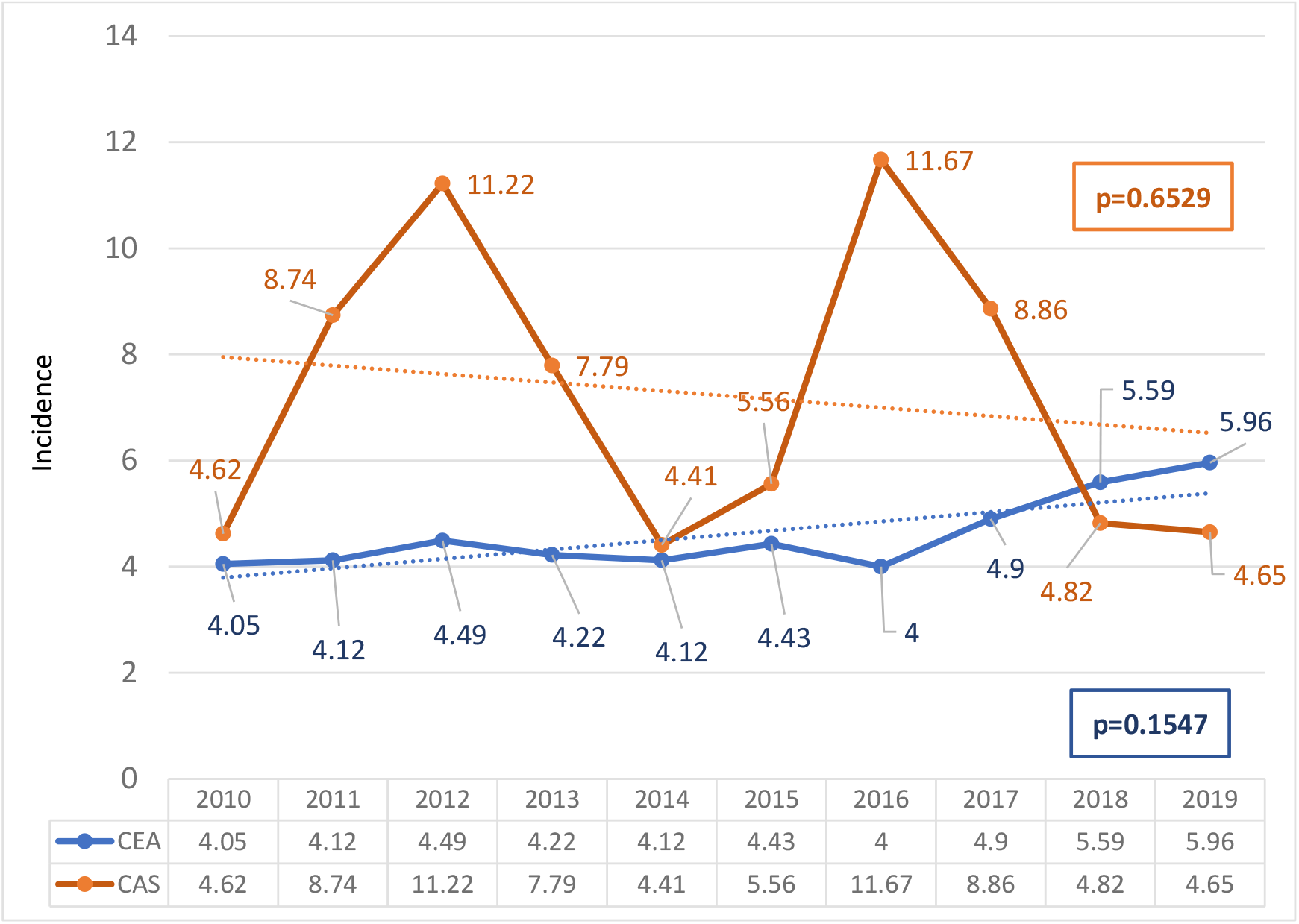
30-day post discharge incidence of percentage of stroke or death following carotid revascularization performed in asymptomatic ESKD patients from 2010-2019. CEA: Carotid Endarterectomy, CAS: Carotid Artery Stenting

### In-Hospital and One-Year Outcomes

In-hospital stroke or death occurred in 3.39% of CAS patients and 2.22% of CEA patients (P=0.0433). At one year, the composite outcome of stroke or death was higher in CAS patients (33.1%) than in the CEA patients (25.4%) (P<0.001), primarily driven by higher all-cause mortality (30.2% for CAS vs 23.2% for CEA) (Figures 4 and 5). There was no significant temporal change in the one-year stroke or death rates for both procedures (P for trend: CEA=0.85; CAS=0.15).

**Figure 4:**
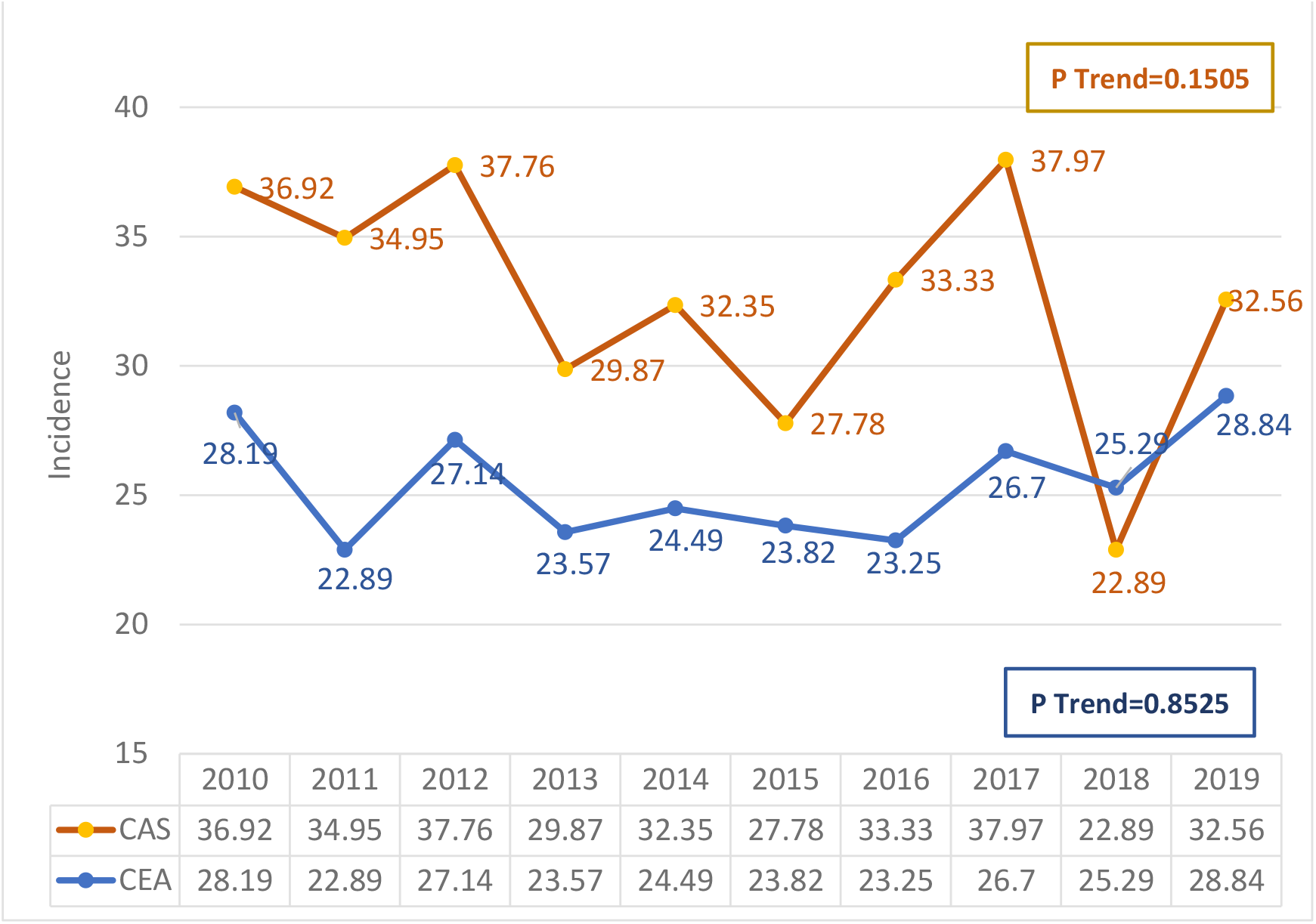
Trends of 1-year post discharge incidence of stroke or death following carotid revascularization performed in asymptomatic ESKD patients from 2010-2019. CEA: Carotid Endarterectomy, CAS: Carotid Artery Stenting

**Figure 5:**
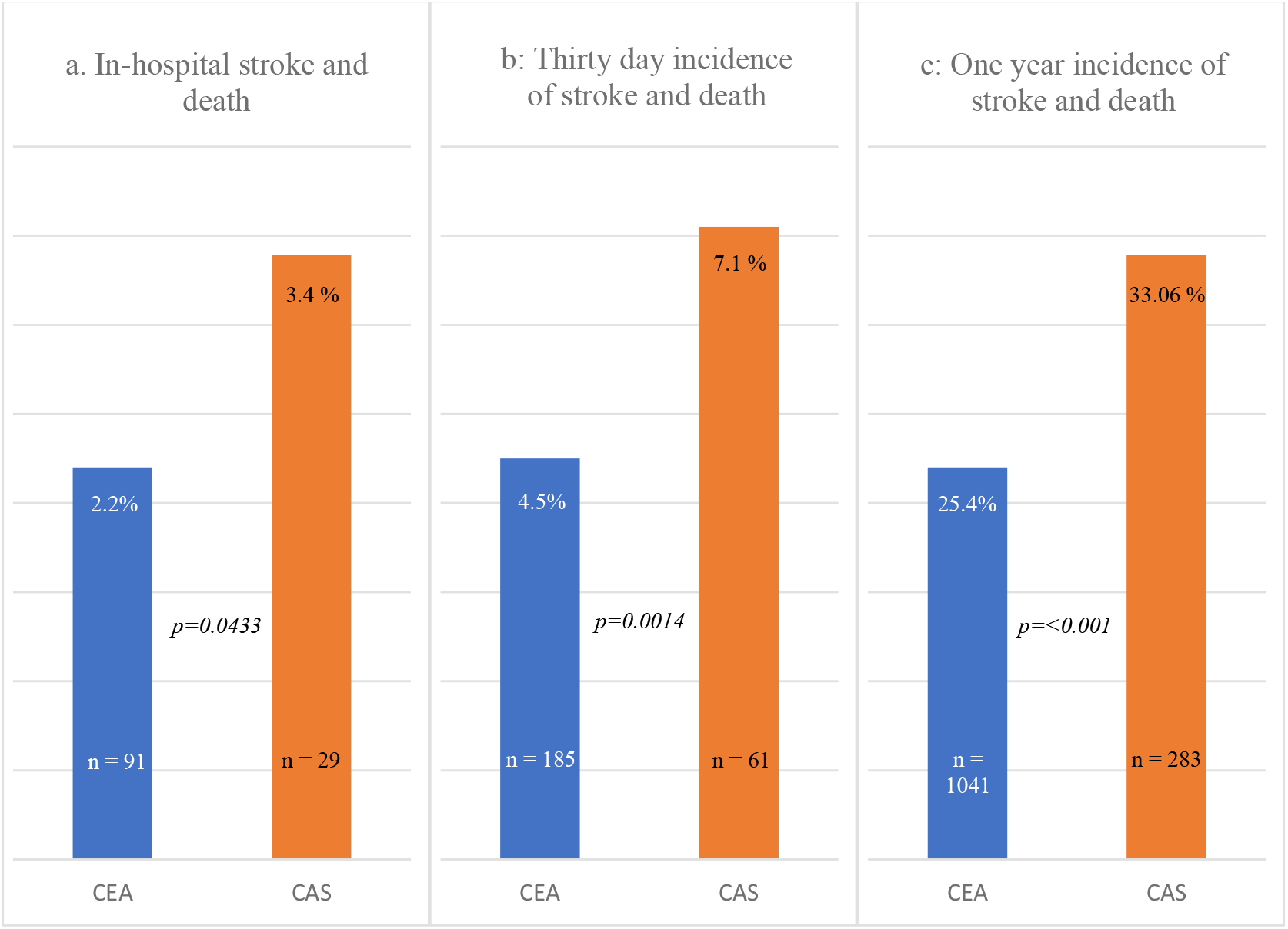
Overall outcomes following revascularization performed for asymptomatic carotid disease in ESKD patients on dialysis from 2010-2019. CEA: Carotid Endarterectomy, CAS: Carotid Artery StentinG

## DISCUSSION

In this national observational cohort of patients with ESKD on dialysis and ACAS, we found that utilization of carotid revascularization declined significantly from 2010 to 2019, with sharper reductions observed in CEA compared to CAS. Despite these declines, a notable proportion of patients still continue to undergo these procedures. Both CEA and CAS were associated with high short-term and one-year rates of stroke or death.

These findings are consistent with the already prevalent concerns about the net clinical benefit of carotid revascularization in ESKD patients, a population with high mortality and known to have a high risk of adverse periprocedural outcomes. Prior randomized clinical trials have shown a modest reduction in the stroke risk with carotid revascularization in asymptomatic patients with severe carotid stenosis (3-5, 20, 21), most of them excluded or included only a small number of dialysis-dependent patients. Our results confirm earlier observational studies indicating poor outcomes following revascularization in this group (9-16).

In the current analysis, 30-day stroke or death was experienced by 4.5% of patients with CEA and 7.1% of patients with CAS, clearly exceeding the <3% periprocedural stroke/death threshold recommended by most clinical guidelines for offering revascularization to ACAS patients with limited comorbidity (6, 7). Moreover, the one-year mortality was approaching 30% in the CAS group and 23% in the CEA group, with no evidence of improvement over time. These findings raise concerns about the appropriateness of carotid revascularization in dialysis-dependent ACAS patients who may not survive long enough to receive any benefit from these stroke prevention procedures.

Our findings are consistent with prior analysis from the USRDS, where You et al. (9) reported a 30-day stroke/death rate of about 10% following CEA or CAS among dialysis patients with ACAS and concluded to avoid revascularization in this population. Similarly, Cooper et al. (11) showed a poor periprocedural and long-term survival after CEA in dialysis patients, including asymptomatic patients. Although our observed complication rates were lower than these prior studies, they remain higher than the guideline thresholds, and no significant improvements in the outcomes were observed during the study period.

The consistent reduction in procedure rates observed in our study may reflect a growing recognition of these risks and significant improvements in the optimal medical therapy for ACAS. Recent advances in medical treatment, including antiplatelet therapy, statins, stringent glucose control, blood pressure control, and tobacco cessation and lifestyle modifications, have narrowed the absolute benefit of revascularization procedures even in the relatively healthier population (22, 23). Nonetheless, about 5,000 EKSD patients still underwent CEA or CAS during the study period. The reason for the continued offering of revascularization is unclear but may relate to provider practice patterns, unclear symptom assessment, and/or extrapolation of general population data. Further efforts to develop and disseminate ESKD-specific guidelines are warranted.

## LIMITATIONS

Our analysis of the administrative database may be subject to coding errors and unable to capture the laterality or anatomical severity of the carotid disease. Although we employed a 6-month lead-in period to define asymptomatic disease, this approach cannot completely eliminate residual misclassification. We did not have detailed information on medical therapy, procedure technique, and the interventionalist’s experience. Despite our sincere efforts to demonstrate associations between procedure type and outcomes, unmeasured confounding may always persist. Finally, the results may not be generalizable to non-US populations or those without Medicare coverage.

## CONCLUSION

In a national cohort of dialysis-dependent ESKD patients with ACAS, the rates of carotid revascularization have declined over the past decade. Both CEA and CAS were associated with high short-term and one-year rates of stroke or death, with no evidence of improvements over time. Our findings suggest a cautious approach is prudent for revascularization in patients with ESKD and underscore the need for individualized, risk-based shared decision-making in managing asymptomatic carotid disease.

## Data Availability

USRDS Database

https://www.niddk.nih.gov/about-niddk/strategic-plans-reports/usrds

## ABBREVIATIONS

BMI: Body Mass Index
CABG: Coronary Artery Bypass Grafting
CAD: Coronary Artery Disease
CAS: Carotid Artery Stenting
CEA: Carotid Endarterectomy
CHF: Congestive Heart Failure
CKD: Chronic Kidney Disease
CMS: Centers for Medicare & Medicaid Services
COPD: Chronic Obstructive Pulmonary Disease
CVA: Cerebrovascular Accident
DM: Diabetes Mellitus
ESKD: End-Stage Kidney Disease
HDL: High-Density Lipoprotein
ICD: International Classification of Diseases
LDL: Low-Density Lipoprotein
MI: Myocardial Infarction
NSQIP: National Surgical Quality Improvement Program
PVD: Peripheral Vascular Disease
TIA: Transient Ischemic Attack

## REFERENCES

1. Centers for Disease Control and Prevention (CDC). National Vital Statistics System – Mortality (NVSS-M). https://health.gov/healthypeople/objectives-and-data/data-sources-and-methods/data-sources/national-vital-statistics-system-mortality-nvss-m. Accessed June 5, 2025.

2. Yahalom G, Schwartz R, Schwammenthal Y, et al. Chronic kidney disease and clinical outcome in patients with acute stroke. Stroke. 2009;40(4):1296–1303.

3. Ederle J, Dobson J, Featherstone RL, et al. Carotid artery stenting compared with endarterectomy in patients with symptomatic carotid stenosis (International Carotid Stenting Study): an interim analysis of a randomized controlled trial. Lancet. 2010;375:985–997.

4. Tsukahara T. Surgical treatment of carotid artery stenosis. Acta Neurochir Suppl. 2016;123:109–114.

5. Ferguson GG, Eliasziw M, Barr HW, et al. The North American Symptomatic Carotid Endarterectomy Trial: surgical results in 1415 patients. Stroke. 1999;30:1751–1758.

6. Naylor R, Rantner B, Ancetti S, et al. European Society for Vascular Surgery (ESVS) 2023 clinical practice guidelines on the management of atherosclerotic carotid and vertebral artery disease. Eur J Vasc Endovasc Surg. 2022;65:7–111.

7. AbuRahma AF, Avgerinos ED, Chang RW, et al. Society for Vascular Surgery clinical practice guidelines for management of extracranial cerebrovascular disease. J Vasc Surg. 2022;75(1S):4S–22S.

8. Brott TG, Halperin JL, Abbara S, et al. 2011 ASA/ACCF/AHA/AANN/AANS/ACR/ASNR/CNS/SAIP/SCAI/SIR/SNIS/SVM/SVS guideline on the management of patients with extracranial carotid and vertebral artery disease. Circulation. 2011;124:e489–e532.

9. You TH, Sidaoui J, Marone LK, et al. Revascularization of asymptomatic carotid stenosis is not appropriate in patients on dialysis. J Vasc Surg. 2015;61(3):670–674.

10. Hafeez MS, et al. Carotid endarterectomy should not be recommended to end-stage kidney disease patients with asymptomatic carotid artery disease. Ann Vasc Surg. 2024;101:53–61.

11. Cooper M, Arhuidese IJ, Obeid T, et al. Perioperative and long-term outcomes after carotid endarterectomy in hemodialysis patients. JAMA Surg. 2016;151(10):947–952.

12. Sidawy AN, Aidinian G, Johnson ON III, White PW, DeZee KJ, Henderson WG. Effect of chronic renal insufficiency on outcomes of carotid endarterectomy. J Vasc Surg. 2008;48(6):1423–1430.

13. Rigdon EE, Monajjem N, Rhodes RS. Is carotid endarterectomy justified in patients with severe chronic renal insufficiency? Ann Vasc Surg. 1997;11(2):115–119.

14. Reil T, Shekherdimian S, Golchet P, Moore W. The safety of carotid endarterectomy in patients with preoperative renal dysfunction. Ann Vasc Surg. 2002;16(2):176–180.

15. Debing E, Van den Brande P. Chronic renal insufficiency and risk of early mortality in patients undergoing carotid endarterectomy. Ann Vasc Surg. 2006;20(5):609–613.

16. Avgerinos ED, Go C, Ling J, Makaroun MS, Chaer RA. Survival and long-term cardiovascular outcomes after carotid endarterectomy in patients with chronic renal insufficiency. Ann Vasc Surg. 2015;29(1):15–21.

17. United States Renal Data System. 2012 Researcher’s Guide to the USRDS Database. http://www.usrds.org/2012/rg/A_intro_sec_1_12.pdf. Accessed June 5, 2025.

18. Saran R, Robinson B, Abbott KC, et al. US Renal Data System 2016 Annual Data Report: Epidemiology of kidney disease in the United States. Am J Kidney Dis. 2017;69(3)(suppl 1):Svii–Sviii.

19. U.S. Department of Health & Human Services. Health information privacy: research. http://www.hhs.gov/ocr/privacy/hipaa/understanding/coveredentities/research.html. Accessed June 5, 2025.

20. Ferguson GG, Eliasziw M, Barr HW, et al. The North American Symptomatic Carotid Endarterectomy Trial: surgical results in 1415 patients. Stroke. 1999;30:1751–1758.

21. Brott TG, Hobson RW, Howard G, et al. Stenting versus endarterectomy for treatment of carotid-artery stenosis. N Engl J Med. 2010;363:11–23.

22. Meschia JF, Bushnell C, Boden-Albala B, et al. Guidelines for the primary prevention of stroke: a statement for healthcare professionals from the American Heart Association/American Stroke Association. Stroke. 2014;45:3754–3832.

23. Wang T, Sun X, Wang X, et al. Carotid revascularisation versus medical treatment for asymptomatic carotid artery stenosis. Cochrane Database Syst Rev. 2024;7(7):|pCD015499. doi:10.1002/14651858.CD015499.

